# Epigenetic prediction of major depressive disorder

**DOI:** 10.1101/19001123

**Authors:** Miruna C. Barbu, Rosie M. Walker, David M. Howard, Kathryn L. Evans, Heather C. Whalley, David J. Porteous, Stewart W. Morris, Ian J. Deary, Riccardo E. Marioni, Toni-Kim Clarke, Andrew M. McIntosh

## Abstract

**Objective:** DNA methylation (DNAm) is associated with environmental risk factors for major depressive disorder (MDD) but has not yet been tested for its ability to discriminate individuals with MDD from unaffected individuals.

**Methods:** Using penalized regression based on genome-wide CpG methylation, we trained a DNAm risk score of MDD (DNAm-RS) in 1,223 cases and 1,824 controls and tested in a second independent sample of 363 prevalent cases and 1,417 controls. Using DNA from 1,607 unaffected individuals, we tested whether DNAm-RS could discriminate the 190 incident cases of lifetime MDD from the 1,417 individuals who remained unaffected at follow-up.

**Results:** A weighted linear combination of 196 CpG sites were derived from the training sample to form a DNAm-RS. The DNAm-RS explained 1.75% of the variance in MDD risk in an independent case-control sample and significantly predicted future incident episodes of MDD at follow up (R^2^=0.52%). DNAm-RS and MDD polygenic risk scores together additively explained 3.99% of the variance in prevalent MDD. The DNAm-RS was also significantly associated with lifestyle factors associated with MDD, including smoking status (β=0.440, p=<2×10^−16^) and alcohol use (β=0.092, p=9.85×10^−5^). The DNAm-RS remained significantly associated with MDD after adjustment for these environmental factors (independent association: β=0.338, p=1.17×10^−7^ association post-adjustment: β=0.081, p=0.0006).

**Conclusions:** A novel risk score of MDD based on DNAm data significantly discriminated MDD cases from controls in an independent dataset, and controls who would subsequently develop MDD from those who remained unaffected. DNAm-RS captured the effects of exposure to key lifestyle risk factors for MDD, revealing a potential role in risk stratification.

## Introduction

Major Depressive Disorder (MDD) is a frequently disabling condition with an estimated point prevalence of 4.4% (1). Recent genome-wide association studies (GWASs) have begun to elucidate the genetic architecture of MDD (2; 3) and polygenic risk scores (PRS) derived from the most recent study explain 1.5-3.2% of MDD risk in independent cohorts (4). As sole predictors of MDD status, PRS have limited clinical utility and may not capture the larger environmental contributions to risk.

Variation in DNA methylation (DNAm) in affected by both of genetic and environmental factors, which act in combination to confer risk for diseases and complex traits (5), and has recently been studied in relation to MDD. An epigenome-wide association study (EWAS) of 7,948 European individuals identified 3 CpG sites that were differentially methylated in association with depressive symptoms (6). Annotation of these sites implicated genes involved in axon guidance. A study of 150 MZ twins discordant for early-onset MDD identified 760 differentially methylated CpG sites which mapped to neuronal circuitry and plasticity genes (7). These findings suggest that differences in DNAm may be relevant to the causes of MDD.

Many lifestyle factors associated with MDD are also robustly associated with DNAm. Smoking (8), obesity (9; 10) and alcohol consumption (11) are each associated with differential genome-wide DNAm. These DNAm signatures have been leveraged, using penalized regression to identify a subset of informative CpG sites, to create DNAm risk scores (DNAm-RS) which can predict the trait of interest in an independent cohort. McCartney and colleagues showed that DNAm scores explained 61% of the variance in smoking status and 12.5% of the variance in body mass index (BMI) and alcohol consumption. When modelled alongside PRS, DNAm scores contribute additively to the variance explained for these traits (12). DNAm therefore acts as an archive of exposure to several risk factors for poor mental health, but its value as a predictor of MDD, however, remains unexplored.

The aim of this study was to use penalized regression to train a predictor of MDD based on DNAm in the Generation Scotland: Scottish Family Health Study (GS:SFHS) cohort. A training set of 1,223 MDD cases and 1,824 controls was used to create an MDD DNAm risk score (DNAm-RS) in 1,970 independent individuals (363 prevalent and 190 incident cases; 1,417 controls). Using longitudinal clinical data, we also tested whether baseline DNAm-RS would predict future MDD status at follow-up between 4-10 years later. Finally, to explore whether the MDD DNAm-RS captures exposure to lifestyle factors associated with MDD, we tested the association between MDD DNAm-RS and alcohol use, BMI, smoking status, and pack years, as well as self-reported antidepressant use.

## Methods

### Study population: *Generation Scotland - the Scottish Family Health Study (GS:SFHS)*

Phenotypic information, DNAm data and genotypes were provided by GS:SFHS for the current investigation. GS:SFHS is a family-based population cohort investigating the genetics of health and disease in approximately 24,000 individuals across Scotland (13; 14). Baseline data were collected between 2006 and 2011. The present study focuses on 5,017 individuals for whom DNAm data from a blood draw at baseline contact, baseline phenotypic data, and genotype data were available. Environmental data, such as lifestyle factors, were also measured (BMI) or recorded (smoking status, alcohol consumption) on nearly all study participants.

Longitudinal phenotypic data is available for a subset of individuals who responded to a recontact request. For these individuals we have information on MDD case-control status both at baseline and follow-up, which occurred 4-10 years later (2015-2016). GS:SFHS received ethical approval from NHS Tayside Research Ethics Committee (REC reference number 05/S1401/89) and has Research Tissue Bank Status (reference: 15/ES/0040). W

### Phenotypes

BMI was calculated using height (cm) and weight (kg) measured by clinical staff during baseline recruitment. Alcohol intake was self-reported as part of a pre-clinical questionnaire. Participants were asked whether they were ‘never’ ‘former’ or ‘current’ drinkers. Current drinkers were asked: “During the past week, please record how many units of alcohol you have had”. Smoking status was recorded by asking participants: “Have you ever smoked tobacco?” Answers were recorded as: “Yes, currently smoke; Yes, but stopped within the past 12 months; Yes, but stopped more than 12 months ago; No, never smoked”. For the current study, we assigned smoking status as a binary variable, by converting all “Yes” answers to smoker (1), and “No” to non-smoker (0). Using smoking behaviour data, pack years were calculated by multiplying the number of cigarette packs (20 cigarettes/pack) smoked per day by the number of years a person has smoked (16). Antidepressant use was self-reported by participants at the baseline assessment and has been described in greater detail previously (17; Supplementary Material).

Baseline MDD status was measured using the axis-I Structured Clinical Interview of the Diagnostic and Statistical Manual, version IV (SCID) and was administered to participants who answered “yes” to either of two screening questions (see supplemental methods). MDD status was measured prospectively by remote paper questionnaire between 4 and 10 years after baseline assessment (2015-2016) using the Composite International Diagnostic Interview - Short Form (CIDI-SF) as described previously (15).

Control participants were defined as those individuals who answered “no” to the two screening questions (see supplemental methods) and did not fulfill criteria for a diagnosis of current or previous MDD following the SCID interview and CIDI-SF remote follow-up assessment. Individuals fulfilling criteria for schizophrenia or bipolar disorder, or who self-reported these diagnoses, were also excluded from both case and control groups.

### DNA methylation

9,873 individuals in GS:SFHS had genome-wide DNAm data profiled from blood samples using the Illumina Human-MethylationEPIC BeadChip. The raw data were acquired, preprocessed and quality checked in two different batches, hereafter named batch 1 (N = 5,190) and batch 2 (N = 4,588).

In batch 1, ShinyMethyl (18) was used to exclude samples where predicted sex mismatched recorded sex, as well as to plot the log median intensity of methylated and unmethylated signals per array and inspect the output from the control probes; outlying samples detected by visually inspection were excluded. WateRmelon (19) was then used to remove probes in which > 1% of cytosine-guanine dinucleotide had a detection p-value > 0.05; probes with a beadcount of < 3 in more than 5% samples; and probes in which > 5% of samples had a detection p-value > 0.05 (12). Multi-dimensional scaling (MDS) plots were inspected to confirm that there were no additional sample outliers. WateRmelon was then used to normalise the data, data using the dasen method, and lumi (20) was used for conversion to M-values, which were then pre-corrected for relatedness, estimated blood cell types, and processing batch using DISSECT (21), for CpGs on autosomal chromosomes. The final dataset comprised corrected M-values at 841,753 loci measured for 5,087 individuals.

In batch 2, meffil (22) and ShinyMethyl (18) were used for quality control of the raw data. Using meffil, samples were removed if: there was a mismatch between self-reported and methylation-predicted sex; they had > 1% of CpG sites with a detection p-value > 0.05; they showed evidence of dye bias; they were outliers for the bisulphite conversion control probes; and had a median methylated signal intensity > 3 standard deviations lower than expected. Afterwards, shinyMethyl was used to perform further quality control, as described above for batch 1. Multi-dimensional scaling plots were inspected, and outliers were excluded. Meffil was then used again to identify and exclude poor-performing probes, which were deemed as such if: they had a beadcount of < 3 in > 5% samples and/or > 5% samples had a detection p-value > 0.05. The data were normalised using the dasen method in wateRmelon, and the beta2m function in lumi (20) was used to generate M-values. The final dataset comprised M-values for 773,860 loci measured in 4,450 individuals.

### Genotyping and PRS profiling

Individuals were genotyped using the Illumina OmniExpress BeadChip. The raw genotype data underwent a series of quality control steps: individuals with a call rate < 98%, single nucleotide polymorphisms (SNPs) with a genotype rate < 98%, minor allele frequency < 1%, and Hardy-Weinberg p-value < 10^−6^ were removed from the initial dataset and then imputation was performed using the Sanger Imputation Service with the Haplotype Reference Consortium panel v1.1 (23; 4).

Using the largest available depression GWAS (4), depression PRS were computed using Plink v1.90b4 (24) using SNPs that met a significance level of p ≤ 0.05, in line with previous studies which have shown that this threshold explains the most variance in MDD status (4). GWAS summary statistics excluding GS:SFHS were obtained in order to create PRS in the GS:SFHS sample. Clumping was applied using a linkage disequilibrium r^2^ < 0.1 and a 500-kb window.

### DNAm predictor – training and testing datasets

In order to obtain a training and testing dataset, individuals were separated based on the two batches described above. Supplementary Figure 1 provides a flowchart summary of the analysis process.

#### Training dataset

Batch 1 was used to train the DNAm predictor. The dataset consisted of controls who were screened as unaffected (N = 1,824) at both baseline and follow-up (i.e. answered “no” to screening questions at baseline and follow-up), or who screened positive but were subsequently found not to fulfill diagnostic criteria for MDD using the SCID. MDD cases were those who screened positive for depression by answering yes to one or more brief screening questions and who subsequently fulfilled criteria for MDD at baseline SCID interview (N = 1,223). CpG sites measured in these individuals were included as independent variables in a least absolute shrinkage and selection operator (LASSO) penalised regression model described below. Depression status was regressed on age, sex, and ten genetic principal components, and the extracted residuals from this model were input as the dependent variable in the LASSO regression model.

LASSO penalised regression models were run using the “glmnet” function in R in order to train DNAm predictors. We applied tenfold cross-validation and the mixing parameter was set to 1 for our LASSO penalty. 196 CpGs were included in the predictor that corresponded to the minimum mean cross-validated error (see Supplementary Excel file 1 in supplementary materials for a list of CpG sites and their regression weights).

#### Testing dataset

Batch 2 was used in order to create MDD DNAm-RS using the CpG sites identified in the LASSO regression models. To create a single DNAm-RS, the CpG weights corresponding to the 196 CpG sites identified in the training sample were multiplied by the CpG values in the independent sample. The DNAm-RS were tested for association with prevalent MDD cases (depressed at baseline, N=363) and incident MDD cases (healthy at baseline but fulfilling criteria for MDD at follow-up, N=190). The same 1,417 controls were used as the comparison group for both sets of MDD cases and were unaffected at both baseline and follow-up. The incident MDD cases were used to assess if the DNAm-RS could predict a future episode of MDD.

### Statistical methods

#### Association of DNAm-RS with depression

In order to test whether DNAm-RS is associated with prevalent and incident MDD, MDD status was regressed on and (1) DNAm-RS in the prevalent cases and controls (N = 1,780); and (2) DNAm-RS in the incident cases and controls (N = 1,607) using logistic regression. We also tested the association of DNAm-RS with prevalent (N = 1,250) and incident (N = 1,195) MDD in a subset of individuals with no self-reported antidepressant use.

To determine how much phenotypic variance in MDD DNAm-RS explained compared to a genetic PRS, we regressed MDD status on (1) PRS; (2) DNAm-RS; and (3) PRS and DNAm-RS using logistic regression and calculated McFadden’s R^2^ for each variable. In addition, using the “ROCR” R package, we plotted the predictive ability of DNAm-RS in both incident and prevalent cases and controls using a Receiver Operating Characteristic (ROC) curve, representing the sensitivity and specificity of the score in relation to depression.

#### Association of DNAm-RS with lifestyle factors and antidepressant use

We tested whether lifestyle factors previously shown to be associated with both MDD and DNAm (8; 9; 10; 11; 12) were also associated with the DNAm-RS. Using linear regression, we tested whether DNAm-RS associated with BMI, pack years, and alcohol consumption. Logistic regression models were used to test whether DNAm associated with self-reported antidepressant use and smoking status.

#### Association of DNAm-RS with depression when adjusting for lifestyle factors

Finally, to estimate how much variance DNAm-RS explains in MDD status when adjusting for lifestyle factors, MDD status was modelled as a dependent variable with alcohol consumption, BMI, smoking and pack years fit as covariates. We also tested the effect of fitting self-reported antidepressant use in our models to determine whether the DNAm-RS would still significantly contribute to the risk for MDD. This was carried out for both incident and prevalent cases.

## Results

### Association of DNAm-RS with depression

We found DNAm-RS was significantly associated with prevalent (N_total_ = 1,780; cases = 363, controls = 1,417; β = 0.338, p = 1.17×10^−7^, R^2^ = 1.75%) and incident (N_total_ = 1,607; cases = 190, controls = 1,417; β = 0.193, p = 0.016, R^2^ = 0.52%) MDD. After adjustment for self-reported antidepressant use, DNAm-RS was still significantly associated with prevalent MDD (β = 0.236, p = 0.004, R^2^ = 0.77%; independent association: β = 0.338, p = 1.17×10^−7^, R^2^ = 1.75%; Supplementary Table 3). See figure 1 for a ROC curve showing the ability of DNAm-RS to discriminate between MDD cases and controls.

**Figure 1.**
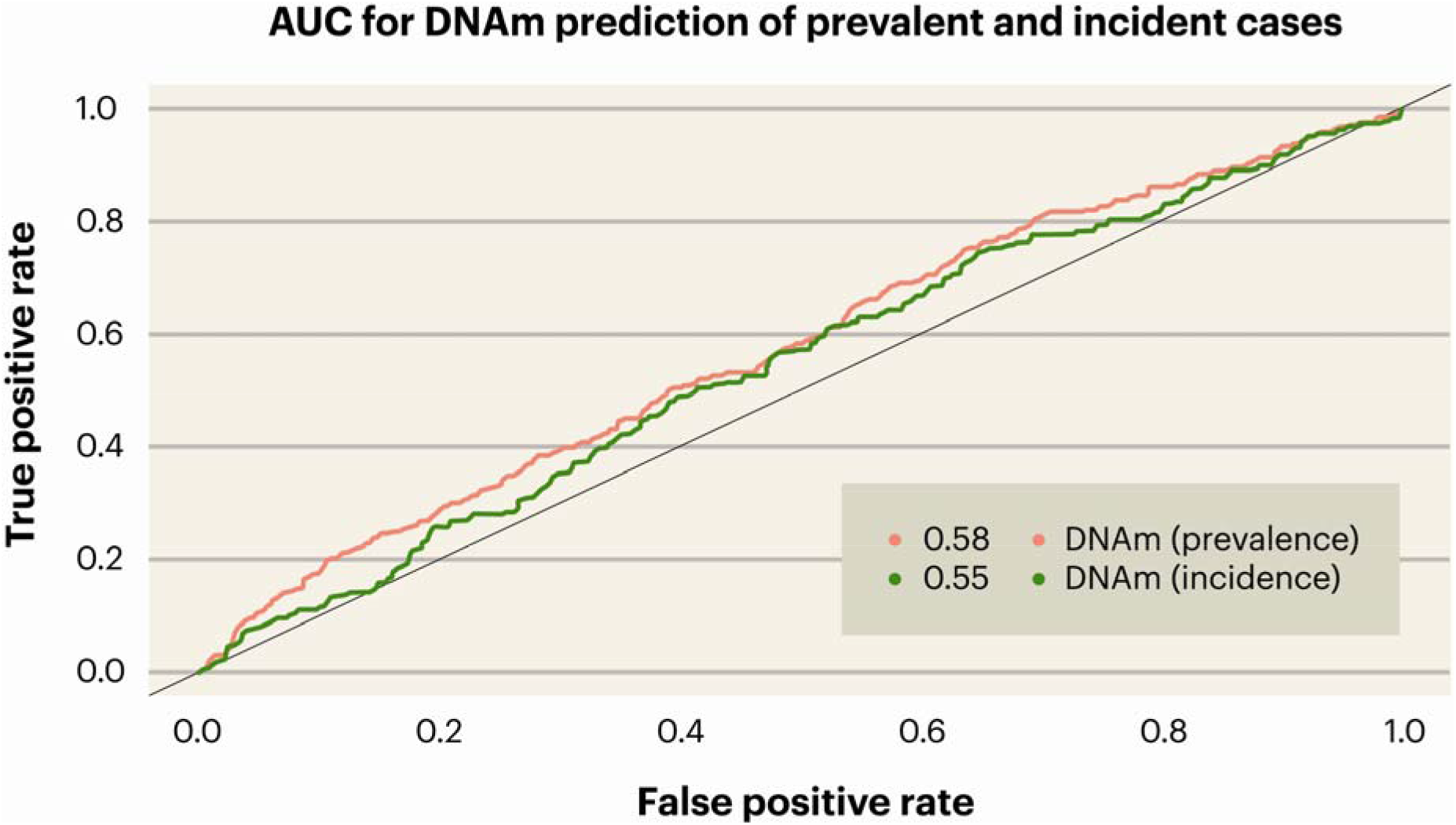
Receiver Operating Characteristic (ROC) curve indicating the sensitivity and specificity of DNAm-RS for both prevalent and incident MDD. The legend shows the AUC estimates for DNAm-RS.

Both DNAm-RS (β = 0.338, p = 1.17×10^−7^, R^2^ = 1.75%) and PRS (β = 0.397, p=1.02×10^−9^, R^2^ = 2.40%) accounted for a small proportion of the variance in risk of prevalent MDD. The model including both DNAm-RS (β = 0.327, p = 5.66×10^−7^) and PRS (β = 0.384, p = 4.69×10^−9^) demonstrated that these two risk scores act additively (R^2^ = 3.99%) and we found no evidence of an interaction (β = −0.009, p = 0.892) (Supplementary Table 1 and Figure 2).

**Figure 2.**
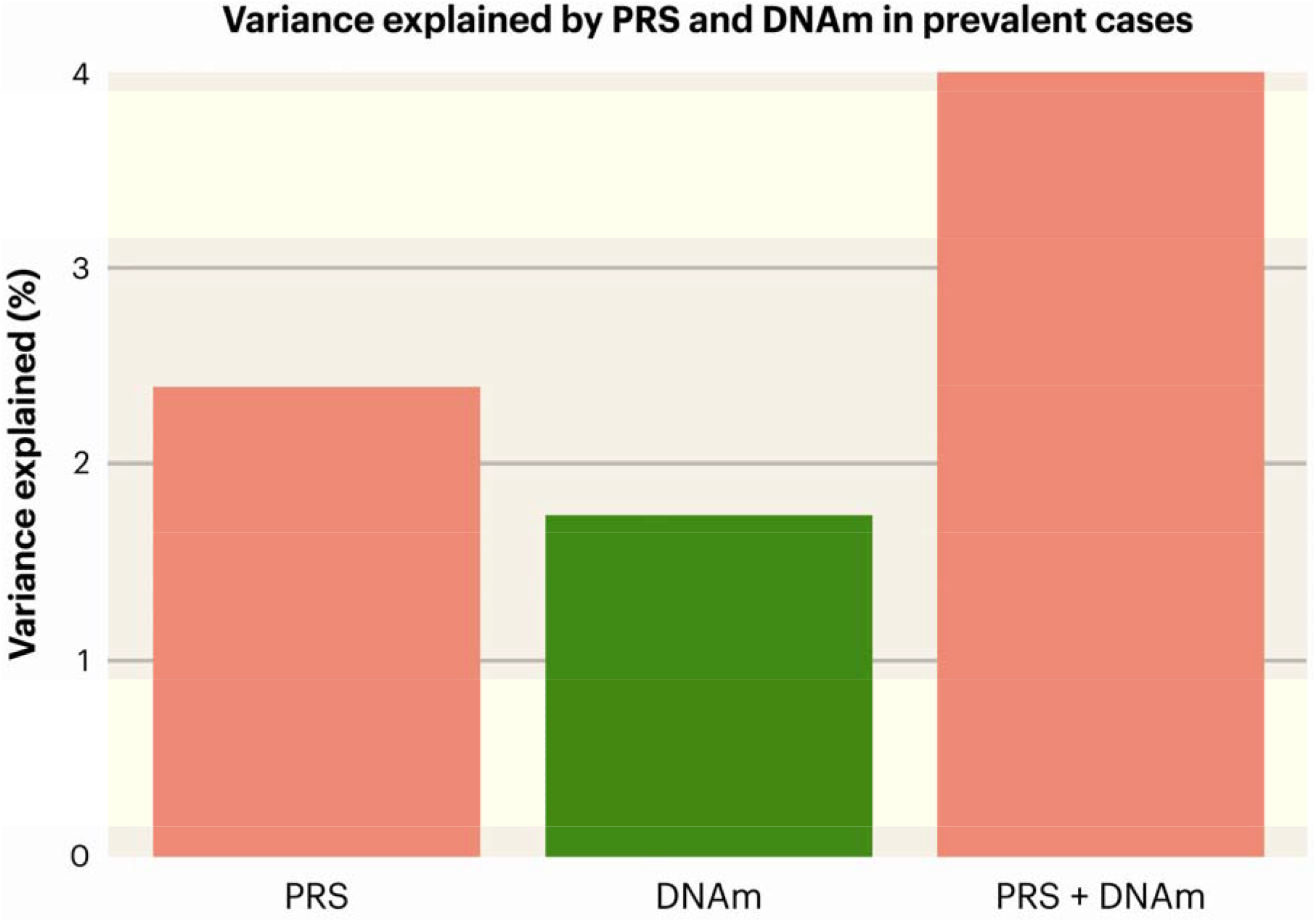
Variance in prevalent MDD (R^2^; y-axis) explained by PRS and DNAm-RS.

### Association of DNAm-RS with depression in cases and controls with no self-reported antidepressant use

In MDD cases and controls with no self-reported antidepressant use (N_Prevalent_ = 1,250, cases = 198, controls = 1,052; N_Incident_ = 1,195, cases = 143, controls = 1,052), DNAm-RS was significantly associated with prevalent (β = 0.331, p = 6.19×10^−5^, R^2^ = 1.66%) and incident (β = 0.232, p = 0.011, R^2^ = 0.76%) MDD. The variance explained in the antidepressant-free subset was slightly lower compared to the full prevalent case-control sample (antidepressant-free sample: R2 = 1.66%; full sample: R^2^ = 1.75%); however, there was no evidence that the DNAm-RS MDD association was significantly attenuated in the antidepressant free incident sub-sample.

### Association of DNAm-RS with lifestyle factors and antidepressant use

DNAm-RS was found to be significantly associated with smoking status (β = 0.440, p = < 2×10^−16^, R^2^ = 3.2%), pack years (β = 0.246, p = < 2×10^−16^, R^2^ = 6.5%), alcohol units (β = 0.092, p = 9.85×10^−5^, R^2^ = 0.7%), and self-reported antidepressant use (β = 0.289, p = 0.002, R^2^ = 1.1%). BMI was not found to be significantly associated with DNAm-RS (β = 0.039, p = 0.099, R^2^ = 0.097%) (Supplementary Table 2; Supplementary Figure 2).

### Association of DNAm-RS with depression when adjusting for lifestyle factors

DNAm-RS was tested for its association with prevalent and incident depression while adjusting for BMI, alcohol use, smoking status and pack years to determine if any independent contribution remained from the DNAm-RS. Table 1 and Figure 3 detail the results. DNAm-RS was still significantly associated with prevalent MDD status after adjusting for lifestyle factors (β = 0.219, p = 0.001) but only explained 0.68% of the variance (independent R^2^ = 1.75%). For incident depression cases, the DNAm-RS was no longer significantly associated with MDD status after adjusting for lifestyle factors (variance explained decreased from 0.52% prior to adjustment to 0.25% after adjustment).

**Table 1.** Standardised effect size, standard error, t value, and nominal p-value for prevalent (P) and incident (I) MDD for lifestyle factors and DNAm-RS.

| Predictor variables | Effect size | SD | t value | p value | R<sup>2</sup> |
| --- | --- | --- | --- | --- | --- |
| **Lifestyle factors + DNAm-RS model (P)** |  |  |  |  |  |
| BMI | 0.259 | 0.061 | 4.247 | $2.16 \times 10^{-5}$ | 1.36% |
| Smoking status | 0.369 | 0.154 | 2.395 | 0.017 | 2.13% |
| Pack years | 0.239 | 0.074 | 3.220 | 0.001 | 1.003% |
| Alcohol units | 0.08 | 0.066 | 1.196 | 0.232 | 0.13% |
| DNAm-RS | 0.219 | 0.067 | 3.259 | 0.001 | 0.68% |
| **Lifestyle factors + DNAm-RS model (I)** |  |  |  |  |  |
| BMI | 0.138 | 0.076 | 1.814 | 0.07 | 0.45% |
| Smoking status | 0.629 | 0.19 | 3.315 | 0.0009 | 1.5% |
| Pack years | -0.0003 | 0.099 | -0.003 | 0.997 | 0.005% |
| Alcohol units | -0.109 | 0.095 | -1.155 | 0.248 | 0.11% |
| DNAm-RS | 0.136 | 0.083 | 1.643 | 0.1 | 0.25% |

**Figure 3.**
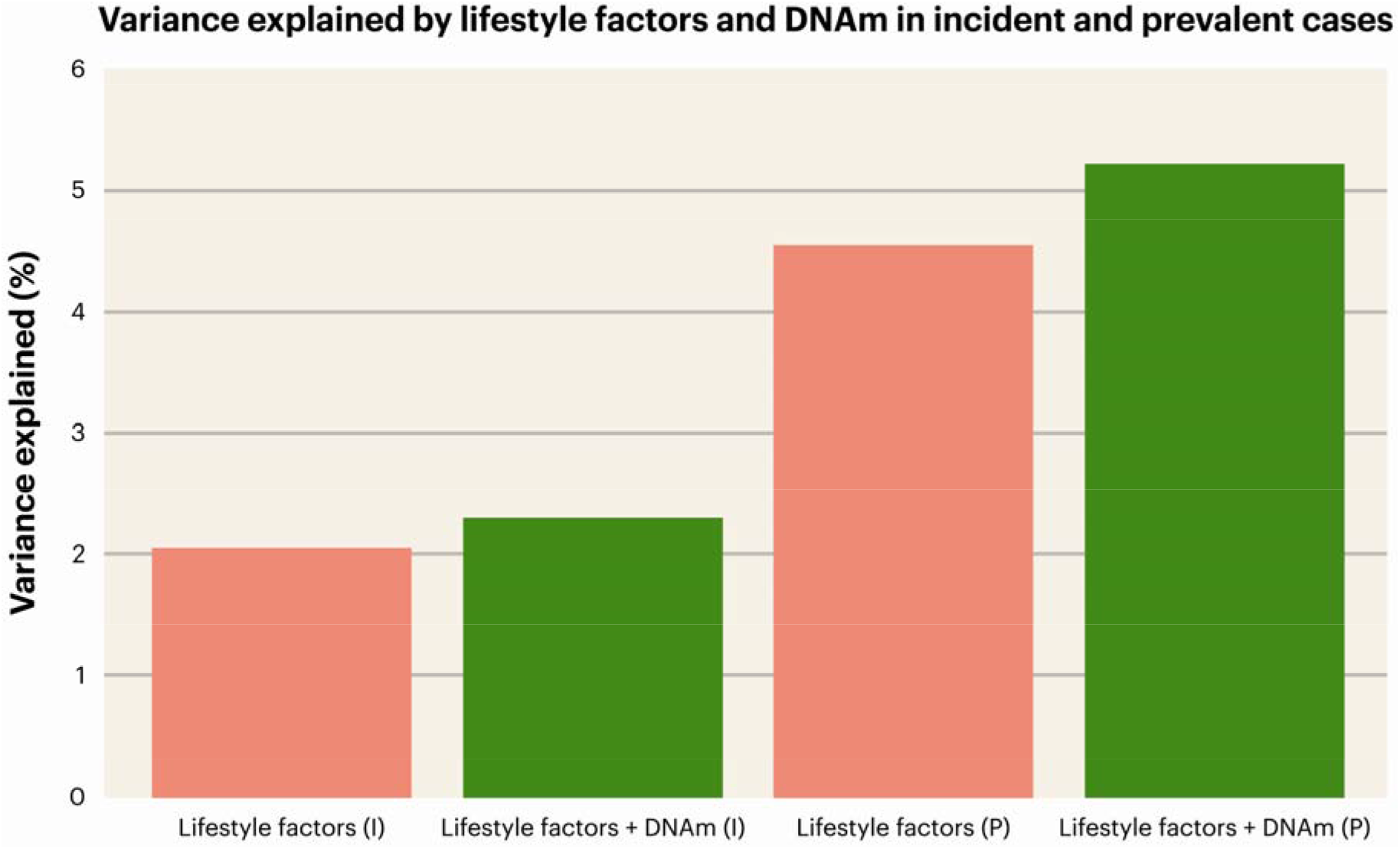
Variance explained (R^2^; y-axis) by lifestyle factors (pink) and lifestyle factors and DNAm-RS (green) in prevalent and incident depression; x-axis labels indicate the independent variables included in the model; I = incidence; P = prevalence.

## Discussion

In the current study we demonstrate that a DNAm-RS explains 1.75% of the variance in MDD status ascertained at the time of blood draw. The genetic PRS explained 2.40% of the variance; additively, the PRS and DNAm-RS account for 3.99% of variance explained in total. Given that MDD PRS scores have been trained on a sample of more than 800K individuals and the DNAm-RS on only 3,047 individuals, the accuracy and clinical potential of DNAm risk scores will likely increase as larger sample sizes with methylation data become available. DNAm risk scores may yet provide clinically-valuable information about the risk of future MDD, albeit that DNAm scores were more weakly associated with future MDD in individuals who were unaffected at baseline, than with case-control status ascertained at the same time as DNA was collected.

In the current study, the MDD DNAm predictor was significantly associated with smoking status and alcohol consumption, but not with BMI (smoking status: R^2^ = 3.2%; alcohol consumption: R^2^ = 0.7%). After adjustment for BMI or smoking status, the DNAm-RS association with MDD was substantially attenuated. These lifestyle factors have previously been associated with MDD (25; 26; 27; 28; 29) and are known to robustly associate with patterns of DNAm (12). The attenuation of the association between DNAm-RS and MDD likely reveals that the DNAm based predictor of MDD may be acting as an archive of the effects of these, and other, lifestyle variables. DNAm-RS was also significantly associated with MDD in a subset of individuals with no self-reported antidepressant use. In addition, the DNAm-RS was also significantly associated with self-reported antidepressant use, although this association does not account for the DNAm-RS MDD associations reported. This finding suggests that DNAm-RS may also be sensitive to the effects of antidepressant use and that future studies should examine whether DNAm-RS trained on antidepressant use may be valuable as a measure of antidepressant absorption or pharmacological action.

The current study, to our knowledge, is the first to investigate a DNAm risk score for MDD in one of the largest samples of DNAm data to date. Using penalised regression models to train our DNAm predictor poses several advantages over other approaches, such as modelling all CpG sites simultaneously or allowing for a non-arbitrary selection of CpG sites, and provided a set of discriminating CpG sites for use in downstream analyses. Moreover, the use of a single score instead of thousands of independent loci allows for a more comprehensive analysis investigating the additive effect of a large number of variants and permits the use of smaller sample sizes. Finally, we were able to gain insight into a novel association between a DNAm-RS and depression, over and above genetic and environmental risk arising from lifestyle factors.

In conclusion, our results show that a DNAm risk score is significantly associated with current and future MDD status, enhancing prediction from polygenic risk scores and environmental traits. Subsequent to further testing and validation in clinically-ascertained samples, these findings may have future clinical applications for MDD risk stratification and justify further efforts to collect DNAm in larger samples.

## Data Availability

The predictor detailed in the manuscript is available in the supplementary Excel file. All other data is available on application to the Generation Scotland Access Committee

## Supplementary materials

### Baseline MDD in GS:SFHS

SCID was administered to participants who answered “yes” to either of the following screening questions: “Have you ever seen anybody for emotional or psychiatric problems?” and “Was there ever a time when you, or someone else, thought you should see someone because of the way you were feeling or acting?” Answers from the SCID were used to ascertain MDD case status.

### Antidepressant use measurement in GS:SFHS

A self-report measure of antidepressant use was recorded by participants in two different ways: within the first phase of the study, a text-based questionnaire was used to record type of antidepressant taken; participants recruited between June 2009 and March 2011 completed a questionnaire recording medication use through a “yes/no” checkbox, with an accompanying question: “Are you regularly taking any of the following medications?”, of which one of the answers was “Antidepressants”.

In the prevalent cases and controls, 108 affected individuals reported they take an antidepressant, whereas 198 reported no antidepressant use; in the incident cases and controls, 20 individuals reported taking an antidepressant, and 143 reported no antidepressant use; 1,052 healthy individuals reported not taking an antidepressant, and 27 report antidepressant use.

---

**Supplementary Figure 1.**
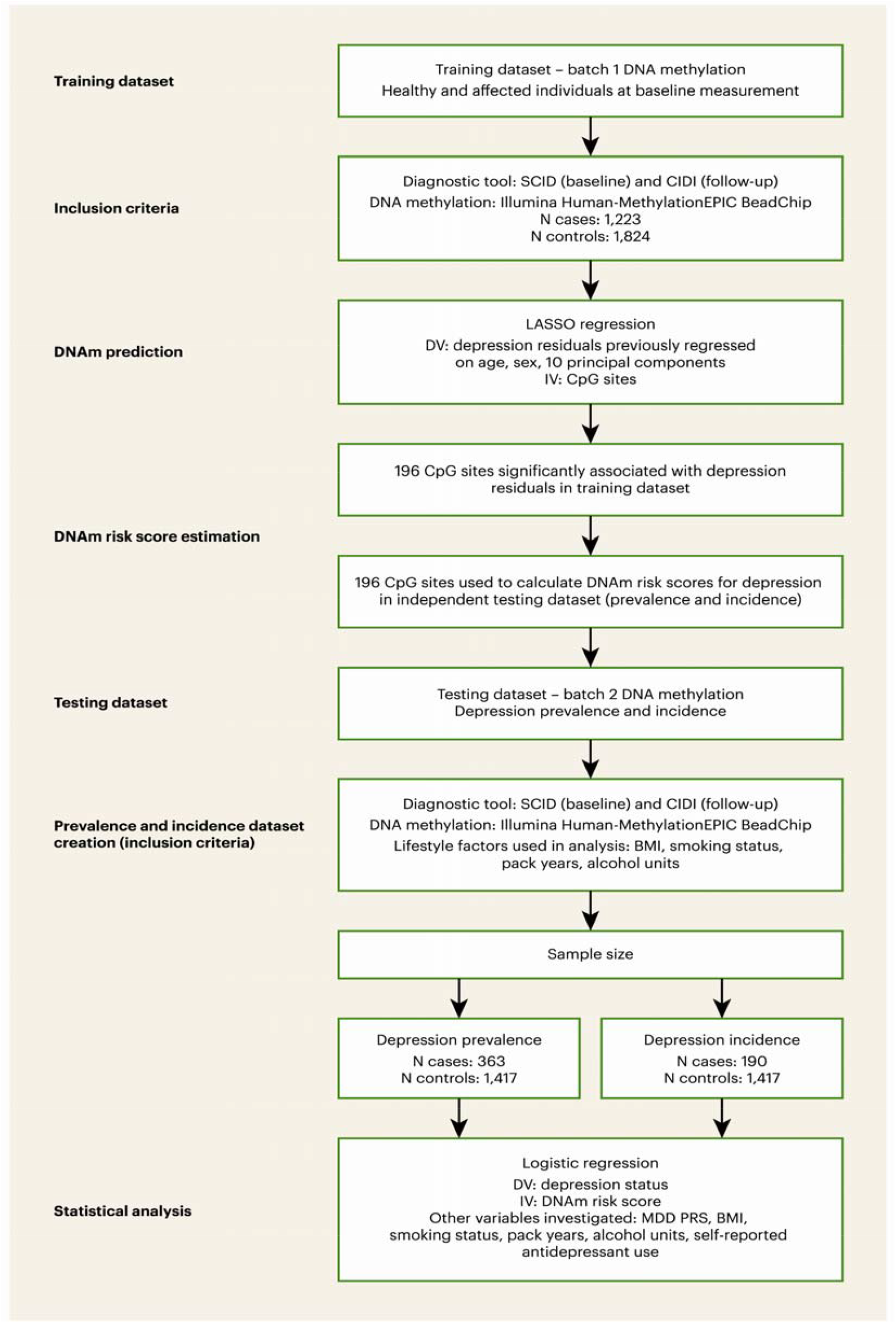
Flowchart indicating analysis process.

**Supplementary Table 1.** Standardised effect size, standard error, t value, nominal p-value, and R^2^ for DNAm, PRS, and PRS + DNAm; IV = independent variable included in the model.

| **Statistic** | **IV: DNAm** | **IV: PRS** | **IV: PRS + DNAm** |
| --- | --- | --- | --- |
| **Effect size, $\beta$** |  |  |  |
| DNAm | 0.338 | - | 0.327 |
| PRS | - | 0.397 | 0.384 |
| **SD** |  |  |  |
| DNAm | 0.064 | - | 0.065 |
| PRS | - | 0.065 | 0.066 |
| **t value** |  |  |  |
| DNAm | 5.289 | - | 5.002 |
| PRS | - | 6.107 | 5.858 |
| **Nominal p value** |  |  |  |
| DNAm | $1.17 \times 10^{-7}$ | - | $5.66 \times 10^{-7}$ |
| PRS | - | $1.02 \times 10^{-9}$ | $4.69 \times 10^{-9}$ |
| **R<sup>2</sup>** | 1.75% | 2.40% | 3.99% |

**Supplementary Table 2.** Standardised effect size, standard error, t value, nominal p-value and R^2^ for lifestyle factors explained by DNAm in prevalent cases and controls (N = 1,780) and self-report antidepressant use (N = 1,385).

| Lifestyle factor | Effect size | SD | t value | p value | R<sup>2</sup> |
| --- | --- | --- | --- | --- | --- |
| BMI | 0.039 | 0.023 | 1.653 | 0.099 | 0.097% |
| Smoking status | 0.440 | 0.051 | 8.598 | $< 2 \times 10^{-16}$ | 3.2% |
| Pack years | 0.246 | 0.022 | 11.197 | $< 2 \times 10^{-16}$ | 6.5% |
| Alcohol units | 0.092 | 0.024 | 3.903 | $9.85 \times 10^{-5}$ | 0.7% |
| Self-report antidepressant use<br>(N = 1385) | 0.289 | 0.091 | 3.165 | 0.002 | 1.1% |

**Supplementary Figure 2.**
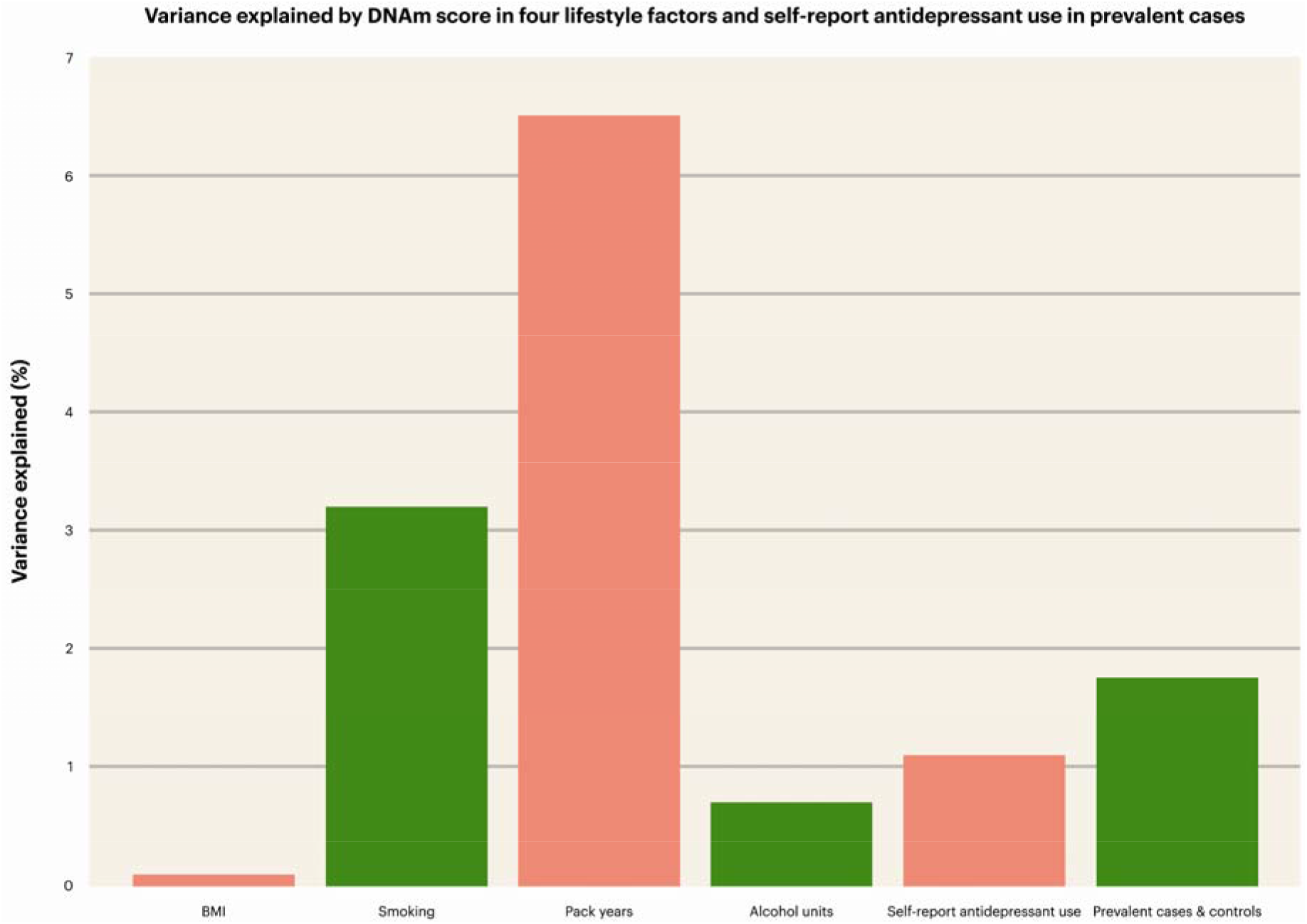
DNAm prediction (R^2^) of lifestyle factors, and prevalent cases & controls (N = 1,780), and self-report antidepressant use (N = 1385).

**Supplementary Table 3.** Standardised effect size, standard error, t value, and nominal p-value for depression explained by lifestyle factors, self-reported antidepressant use, and DNAm in prevalent (P; N = 1,385) and incident (I; N = 1,242) cases and controls; *sample size is lower than in the main manuscript due to self-reported antidepressant use variable.

| Predictor variables | Effect size | SD | t value | p value | R<sup>2</sup> |
| --- | --- | --- | --- | --- | --- |
| **Lifestyle factors + DNAm model (P)** |  |  |  |  |  |
| BMI | 0.197 | 0.077 | 2.553 | 0.011 | 1.62% |
| Smoking status | 0.342 | 0.189 | 1.807 | 0.071 | 2.02% |
| Pack years | 0.111 | 0.953 | 1.166 | 0.244 | 0.62% |
| Alcohol units | 0.124 | 0.079 | 1.566 | 0.117 | 0.11% |
| Self-reported antidepressant use | 3.127 | 0.251 | 12.454 | $< 2 \times 10^{-16}$ | 16.08% |
| DNAm | 0.236 | 0.082 | 2.858 | 0.004 | 0.77% |
| **Lifestyle factors + DNAm model (I)** |  |  |  |  |  |
| BMI | 0.145 | 0.086 | 1.682 | 0.093 | 0.704% |
| Smoking status | 0.624 | 0.211 | 2.963 | 0.003 | 1.66% |
| Pack years | -0.034 | 0.108 | -0.313 | 0.754 | 0.001% |
| Alcohol units | -0.096 | 0.101 | -0.943 | 0.345 | 0.06% |
| Self-reported antidepressant use | 1.603 | 0.33 | 4.857 | $1.19 \times 10^{-6}$ | 2.304% |
| DNAm | 0.143 | 0.092 | 1.549 | 0.121 | 0.27% |

